# A Comprehensive Clinical Description of Pediatric SARS-CoV-2 Infection in Western Pennsylvania

**DOI:** 10.1101/2020.12.14.20248192

**Authors:** Megan Culler Freeman, Kristina Gaietto, Leigh Anne DiCicco, Sherry Rauenswinter, Joseph R. Squire, Zachary Aldewereld, Glenn Rapsinski, Jennifer Iagnemma, Brian T. Campfield, David Wolfson, Traci M. Kazmerski, Erick Forno

## Abstract

**Objective:** We sought to characterize clinical presentation and healthcare utilization for pediatric COVID-19 in Western Pennsylvania (PA).

**Methods:** We established and analyzed a registry of pediatric COVID-19 in Western PA that includes cases in patients <22 years of age cared for by the pediatric quaternary medical center in the area and its associated pediatric primary care network from March 11 through August 20, 2020.

**Results:** Our cohort included 424 pediatric COVID-19 cases (mean age 12.5 years, 47.4% female); 65% reported exposure and 79% presented with symptoms. The most common initial healthcare contact was through telehealth (45%). Most cases were followed as outpatients, but twenty-two patients (4.5%) were hospitalized: 19 with acute COVID-19 disease, and three for multisystem inflammatory syndrome of children (MIS-C). Admitted patients were younger (p<0.001) and more likely to have pre-existing conditions (p<0.001). Black/Hispanic patients were 5.8 times more likely to be hospitalized than white patients (p=0.012). Five patients (1.2%) were admitted to the PICU, including all three MIS-C cases; two required BiPAP and one mechanical ventilation. All patients survived.

**Conclusions:** We provide a comprehensive snapshot of pediatric COVID-19 disease in an area with low to moderate incidence. In this cohort, COVID-19 was generally a mild disease; however, ∼5% of children were hospitalized. Pediatric patients can be critically ill with this infection, including those presenting with MIS-C.

## INTRODUCTION

In December 2019, severe acute respiratory syndrome coronavirus 2 (SARS-CoV-2) emerged as a fatal cause of pneumonia and was quickly recognized as a potentially fatal multisystemic illness termed coronavirus disease 19 (COVID-19)^1^ It has since caused a pandemic of historic proportions. The first confirmed case of SARS-CoV-2 in Allegheny County, in Western Pennsylvania (PA), was on March 11, 2020^2^ While there have been reports on the course of acute COVID-19 in pediatric patients in multiple countries^3–6^ as well as studies detailing multisystem inflammatory syndrome in children (MIS-C)^7–10^ data on pediatric presentation and severity are still limited. Few published reports of COVID-19 include a comprehensive description of pediatric clinical presentation in the United States (US). Most pediatric studies^11^are case reports or series of relatively few patients, often inpatients, with those with over 100 patients reporting numbers of children infected in a region, but not specifically describing their clinical characteristics and disease course.^11^ Many existing studies do not contain child-specific symptom information, often as a result of utilizing ICD code-based retrospective review, and metrics such as underlying medical conditions and hospitalization information are limited.^6,12,13^ In addition, Western PA is representative of a phenomenon seen during the early days of the pandemic, with transition to a model of healthcare delivery utilizing more telehealth and fewer in-person visits^14^ leading to differences in presentation and management of pediatric patients.^14^

From the onset of the pandemic in the region, we created a Pediatric COVID-19 Registry, seeking to characterize the burden of pediatric disease through a collaboration between the University of Pittsburgh Medical Center Children’s Hospital of Pittsburgh (CHP) and the affiliated primary care network, Children’s Community Pediatrics (CCP). CHP is a 315-bed quaternary pediatric care center and admitting hospital that serves a large catchment area, including Western PA, West Virginia, Eastern Ohio, and Western New York. With over 50 locations, CCP is the largest pediatric and adolescent primary care network in Western and Central Pennsylvania. CCP provides care in the Pittsburgh metropolitan area located in Allegheny County, and also in 15 surrounding counties, servicing over 250,000 patients each year. We included all patients receiving care at these facilities who had a confirmed infection with SARS-CoV-2. We then performed a registry-based retrospective chart review to characterize COVID-19 presentation, course of illness, patterns of healthcare utilization, and short-term outcomes.

## METHODS

### Case Inclusion and Data Extraction

At the outset of the pandemic in the region, we created the Western PA Pediatric COVID-19 Registry as a secure RedCap database with the goal of adding cases as they arise during the pandemic and following their course over time^15^ The registry includes all positive SARS-CoV-2 cases under 22 years of age seen at CHP or at CCP via active surveillance since the first reported case in the region; for this analysis, we included cases through August 20, 2020, as this was prior to return-to-school in the region. Subjects are included if they have a positive SARS-CoV-2 RT-PCR or if they meet the case definition for MIS-C. At our institution, criteria for MIS-C aligns with CDC criteria, which include individuals <21 years of age presenting with fever and severe illness requiring hospitalization; laboratory evidence of inflammation (elevated C-reactive protein [CRP], erythrocyte sedimentation rate [ESR], fibrinogen, procalcitonin, d-dimer, ferritin, lactic acid dehydrogenase [LDH], and/or interleukin 6 [IL-6]); multisystem organ involvement; and evidence of recent SARS-CoV-2 infection either by RT-PCR, serology, or antigen test, or known exposure within the four preceding weeks, without other plausible diagnosis. We recorded patient demographics, symptoms, healthcare utilization data, laboratory results, and disease outcomes by reviewing the electronic health records (EHR) at CHP and CCP. If patients were identified both as outpatient and inpatients, only the inpatient presentation was analyzed. Patient information was securely handled as approved by the institutional review board at the University of Pittsburgh (Protocol # STUDY20030250). All data were deidentified prior to analysis.

### Statistical analysis

For cohort description, continuous variables are presented as means and standard deviations unless otherwise stated, while categorical variables are presented as the absolute number in each category and the proportion (percent) of total. We also analyzed demographic and baseline differences between cases requiring hospitalization and those managed as outpatients. We analyzed continuous variables using t-tests and binary/categorical variables using chi-squared tests or ANOVA. We then performed adjusted analyses using logistic regression models to evaluate differences between hospitalized and non-hospitalized patients while adjusting for potential confounders. We considered p-values < 0.05 to be statistically significant. We utilized GraphPad Prism (version 8.0; GraphPad) and STATA v16 (StataCorp LLC) to conduct analyses and used packages choroplethr and choroplethrZip in R v4.0.2 / RStudio v1.3.1073 to produce the choropleth maps.

## RESULTS

From March 11 through August 20, 2020, there were 955 confirmed pediatric cases (age 0-19) of SARS-CoV-2 in Allegheny County, PA, the most populous county in Western PA^2^; 296 (31.0%) of them were included in our registry. During that time, the PA Department of Health (DOH) reported 123,364 total cases in the state, including 10,011 (8.1%) in the 0-19 age group^16^ Counties represented in our registry reported 18,566 total cases during the study period, or 15.0% of the statewide case burden. While several counties do not report age-specific numbers, based on those data we estimate approximately 1,503 pediatric patients in the region during the study period; our registry contains 424 (28.2% of the estimated regional total) and reflects the numbers of pediatric confirmed infections by date (**Figure 1A**). Because most pediatric hospital admissions in the region occur at CHP, our cohort likely includes most hospitalized pediatric cases. **Figure 1B** shows the case distribution by zip code in the Pittsburgh metropolitan area. The distribution of pediatric cases in the registry (**Figures 1D)** mirrors that of total RT-PCR confirmed cases reported by the PA DOH (**Figure 1C)**.

**Figure 1:**
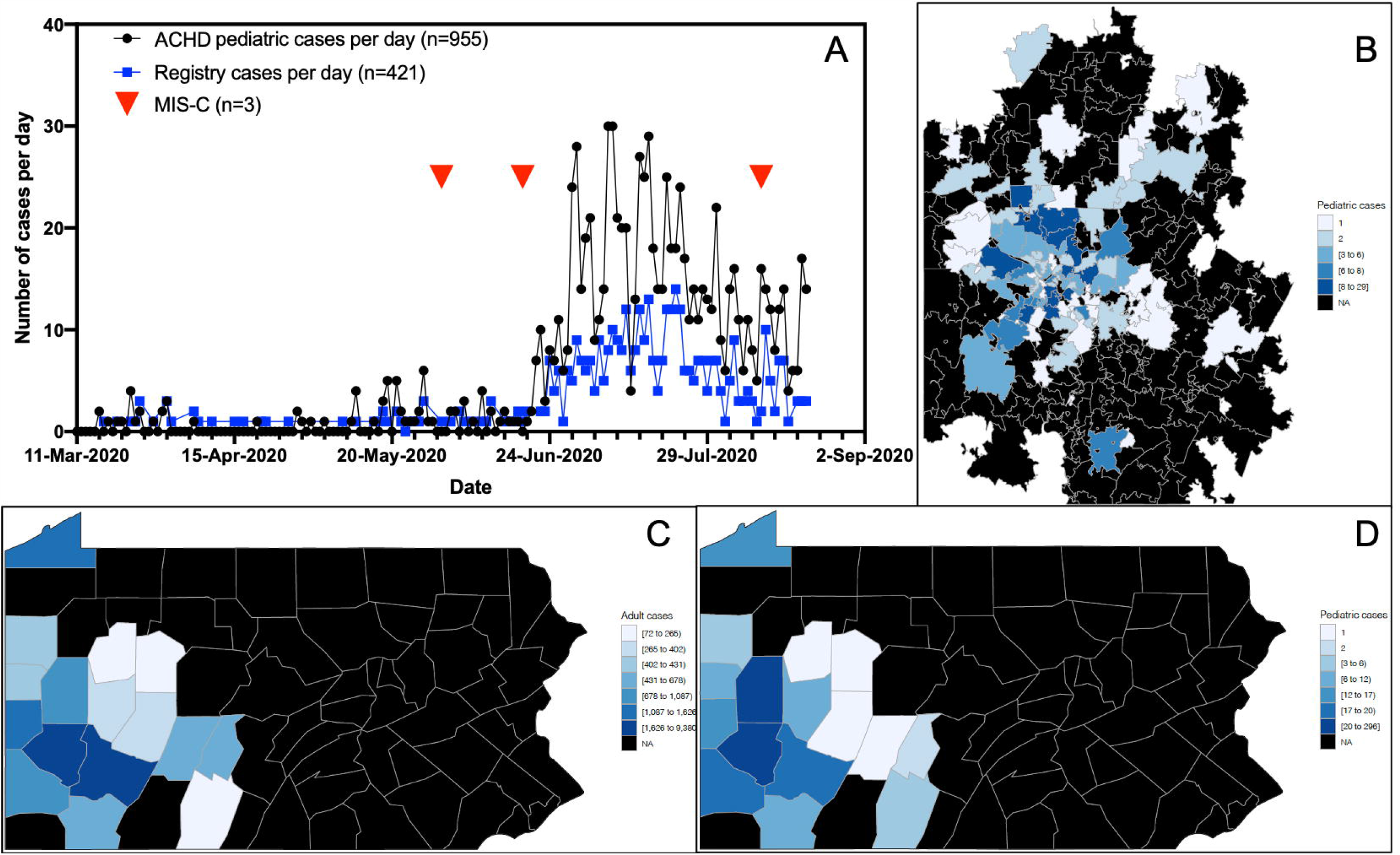
Pediatric COVID-19 cases over time and by geographic location. (A) Allegheny County Health Department (ACHD) total pediatric cases (black) and Registry cases (blue). (B) Pediatric Registry cases by zip code in the Pittsburgh Metropolitan Statistical Area. (C) PA total (adult and pediatric) cases by county. D) Pediatric Registry cases by county, mirroring the distribution of total cases shown in C.

### Demographics and presentation to care

All but three cases in the cohort presented with acute disease (99.3%), while remaining cases presented with MIS-C (0.7%). The mean (SD) age of COVID-19 cases was 12.5 (7.1) years, with a median [IQR] of 15.5 [5.9-18.4] and a range of 7 weeks to 21.9 years (**Table 1, Figure 2**); 52.6% of cases were male. In the cohort, 67.2% of cases identified as white, 24.5% as Black, 3.5% as Asian, and ∼5.2% as other or not reported; six cases (1.4%) identified as Hispanic/Latino. Thirty-seven percent of cases reported a preexisting condition; the most common were asthma (10.6%), neurologic conditions (3.8%), allergic rhinitis (3.5%), cardiac conditions (3.1%), obesity (1.4%), and diabetes (0.9%) (**Table 1, Supplementary Table 1**). For most cases, initial contact with a healthcare provider was either through telehealth visit (45.0%) or phone call to their PCP (26.2%), reflecting the rapid change in practice as a response to the pandemic in our area (**Table 2**). The CHP Emergency Department cared for 19.8% of cases, while 3.3% of cases were identified via CHP preprocedural screening, which began in May 2020.

**Table 1:** Registry Cohort Characteristics.

|  | **Total** | **Outpatient** | **Inpatient** |
| --- | --- | --- | --- |
| Total patients | 424 | 402 (94.8) | 22 (5.2) |
| **Sex** |  |  |  |
| Female | 201 (47.4) | 192 (47.8) | 9 (40.9) |
| Male | 223 (52.6) | 210 (52.2) | 13 (59.1) |
| **Age, years** |  |  |  |
| Mean | 12.5 (7.1) | 12.9 (7.0) | 7.2 (7.4) |
| Median | 15.5 [5.9-18.4] | 15.8 [6.9-18.5] | 5.0 [0.6-13.9] |
| Range (min-max) | 0.1-21.9 | 0.1-21.9 | 0.1-20.9 |
| 0-2 | 79 (18.6) | 69 (17.2) | 10 (45.5) |
| 3-6 | 34 (8.0) | 32 (8.0) | 2 (9.1) |
| 7-12 | 60 (14.2) | 57 (14.2) | 3 (13.6) |
| 13-17 | 117 (27.6) | 113 (28.1) | 4 (18.2) |
| 18-22 | 134 (31.6) | 131 (32.6) | 3 (13.6) |
| **Race / Ethnicity<sup>1</sup>** |  |  |  |
| White | 285 (67.2) | 275 (68.4) | 9 (40.9) |
| Black | 104 (24.5) | 92 (22.9) | 12 (54.5) |
| Asian | 15 (3.5) | 15 (3.7) | 0 (0.0) |
| American Indian/Alaskan Native | 4 (0.9) | 4 (1.0) | 0 (0.0) |
| Hispanic/Latino | 6 (1.4) | 4 (1.0) | 2 (9.1) |
| Other/Not reported | 22 (5.2) | 21 (5.2) | 1 (4.5) |
| **Underlying conditions<sup>2</sup>** |  |  |  |
| Yes<sup>3</sup> | 157 (37.0) | 139 (34.6) | 17 (77.3) |
| - Asthma | 45 (10.6) | 43 (10.7) | 2 (9.1) |
| - Neurologic condition | 16 (3.8) | 12 (3.0) | 4 (18.2) |
| - Allergic rhinitis | 15 (3.5) | 15 (3.7) | 0 (0.0) |
| - Cardiac condition | 13 (3.1) | 11 (2.7) | 2 (9.1) |
| - Obesity | 6 (1.4) | 2 (0.5) | 4 (18.2) |
| - Diabetes | 4 (0.9) | 2 (0.5) | 2 (9.1) |
| - Immunocompromised | 2 (0.5) | 0 (0.0) | 2 (9.1) |
| Chronic home medications | 122 (28.8) | 111 (27.6) | 10 (45.5) |

Data shown as N (%), mean (SD), or median [interquartile range] unless otherwise noted. <sup>1</sup>More than one allowed to be selected per patient. <sup>2</sup>As abstracted from the EHR ICD-10 codes. <sup>3</sup>Number of cases with $\geq 1$ reported chronic condition; patients could have more than one and thus may add up to more than 157.

**Table 2:**
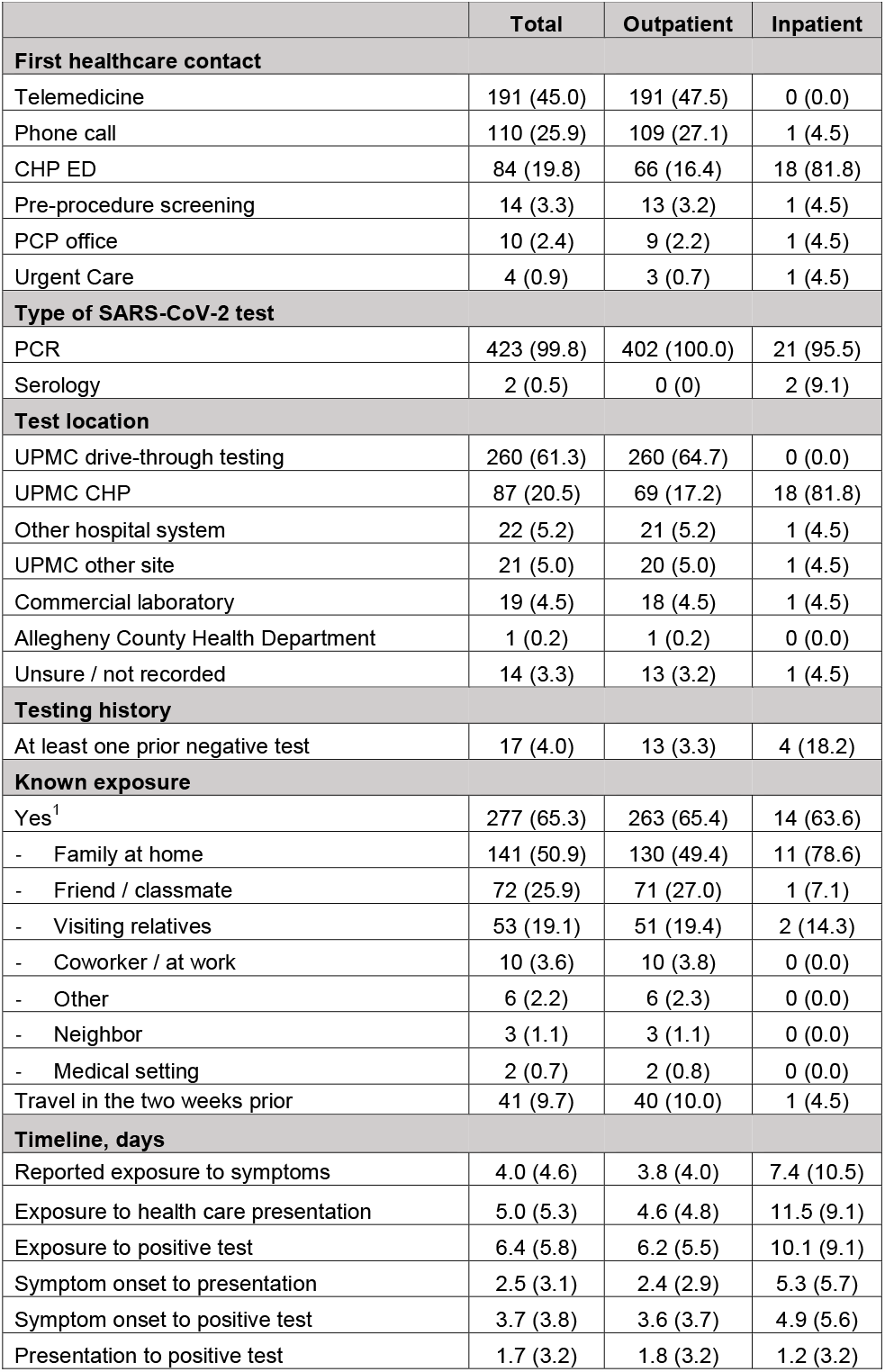
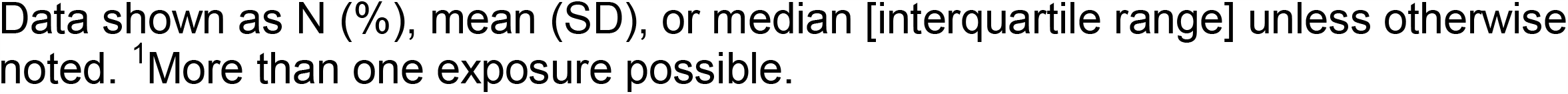
Exposure and Diagnosis.

|  | **Total** | **Outpatient** | **Inpatient** |
| --- | --- | --- | --- |
| **First healthcare contact** |  |  |  |
| Telemedicine | 191 (45.0) | 191 (47.5) | 0 (0.0) |
| Phone call | 110 (25.9) | 109 (27.1) | 1 (4.5) |
| CHP ED | 84 (19.8) | 66 (16.4) | 18 (81.8) |
| Pre-procedure screening | 14 (3.3) | 13 (3.2) | 1 (4.5) |
| PCP office | 10 (2.4) | 9 (2.2) | 1 (4.5) |
| Urgent Care | 4 (0.9) | 3 (0.7) | 1 (4.5) |
| **Type of SARS-CoV-2 test** |  |  |  |
| PCR | 423 (99.8) | 402 (100.0) | 21 (95.5) |
| Serology | 2 (0.5) | 0 (0) | 2 (9.1) |
| **Test location** |  |  |  |
| UPMC drive-through testing | 260 (61.3) | 260 (64.7) | 0 (0.0) |
| UPMC CHP | 87 (20.5) | 69 (17.2) | 18 (81.8) |
| Other hospital system | 22 (5.2) | 21 (5.2) | 1 (4.5) |
| UPMC other site | 21 (5.0) | 20 (5.0) | 1 (4.5) |
| Commercial laboratory | 19 (4.5) | 18 (4.5) | 1 (4.5) |
| Allegheny County Health Department | 1 (0.2) | 1 (0.2) | 0 (0.0) |
| Unsure / not recorded | 14 (3.3) | 13 (3.2) | 1 (4.5) |
| **Testing history** |  |  |  |
| At least one prior negative test | 17 (4.0) | 13 (3.3) | 4 (18.2) |
| **Known exposure** |  |  |  |
| Yes<sup>1</sup> | 277 (65.3) | 263 (65.4) | 14 (63.6) |
| - Family at home | 141 (50.9) | 130 (49.4) | 11 (78.6) |
| - Friend / classmate | 72 (25.9) | 71 (27.0) | 1 (7.1) |
| - Visiting relatives | 53 (19.1) | 51 (19.4) | 2 (14.3) |
| - Coworker / at work | 10 (3.6) | 10 (3.8) | 0 (0.0) |
| - Other | 6 (2.2) | 6 (2.3) | 0 (0.0) |
| - Neighbor | 3 (1.1) | 3 (1.1) | 0 (0.0) |
| - Medical setting | 2 (0.7) | 2 (0.8) | 0 (0.0) |
| Travel in the two weeks prior | 41 (9.7) | 40 (10.0) | 1 (4.5) |
| **Timeline, days** |  |  |  |
| Reported exposure to symptoms | 4.0 (4.6) | 3.8 (4.0) | 7.4 (10.5) |
| Exposure to health care presentation | 5.0 (5.3) | 4.6 (4.8) | 11.5 (9.1) |
| Exposure to positive test | 6.4 (5.8) | 6.2 (5.5) | 10.1 (9.1) |
| Symptom onset to presentation | 2.5 (3.1) | 2.4 (2.9) | 5.3 (5.7) |
| Symptom onset to positive test | 3.7 (3.8) | 3.6 (3.7) | 4.9 (5.6) |
| Presentation to positive test | 1.7 (3.2) | 1.8 (3.2) | 1.2 (3.2) |

Data shown as N (%), mean (SD), or median [interquartile range] unless otherwise noted. <sup>1</sup>More than one exposure possible.

**Figure 2:**
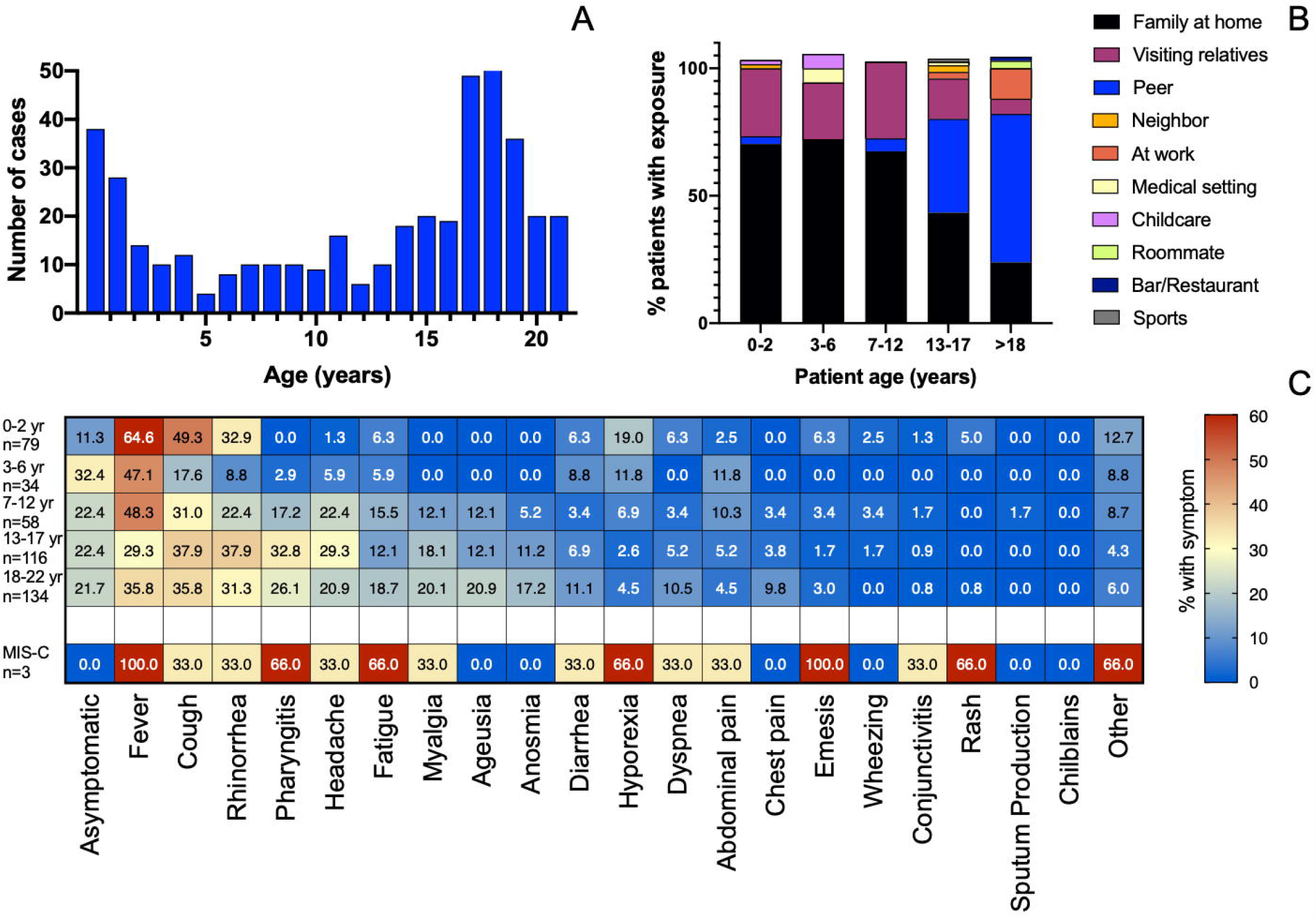
Age distribution, exposures, and symptoms by age group. (A) Histogram of age distribution in pediatric COVID-19 cases. (B) Distribution of exposures by age group. (C) Symptom heatmap by age group, for acute COVID-19 and MIS-C.

### Exposure and testing

Most cases in the cohort tested positive for SARS-CoV-2 via RT-PCR on a nasopharyngeal sample, and two cases of MIS-C had positive IgG serology (**Table 2**). Most testing was performed at the UPMC drive-through testing facility (61.3%), followed by testing at CHP (20.5%). Seventeen cases (4.0%) had previously tested negative for SARS-CoV-2, including four hospitalized patients: two with MIS-C, one with cystic fibrosis, and one who tested negative prior to a neurosurgical procedure, then presented with acute COVID-19 one week later.

Most children had at least one known exposure to another person with COVID-19 (65.3%) (**Table 2**). The most common exposures were other family members at home (50.9%), peers (25.9%), and visiting relatives (19.1%). Exposure varied by age group, with the youngest group (0-2 years) most likely being exposed at home (69.8%), by other family (27.0%), or via childcare (1.6%); and the young adults (18-21 years) exposed by peers (57.4%), at work (11.8%), or at bars/restaurants (1.5%) **(Figure 2B**). Forty-one patients (9.7%) reported travel within the two weeks prior to presentation of illness, mostly to locations requiring air travel.

For cases with available dates of exposure, on average (SD), there were 4 (4.6) days from exposure to symptom onset, 2.5 (3.1) days from symptom onset to health care presentation, and 1.7 (3.2) days from presentation to testing; total interval from symptom onset to testing was 3.7 (3.8) days (**Table 2, Figure 3 A-D**).

**Figure 3:**
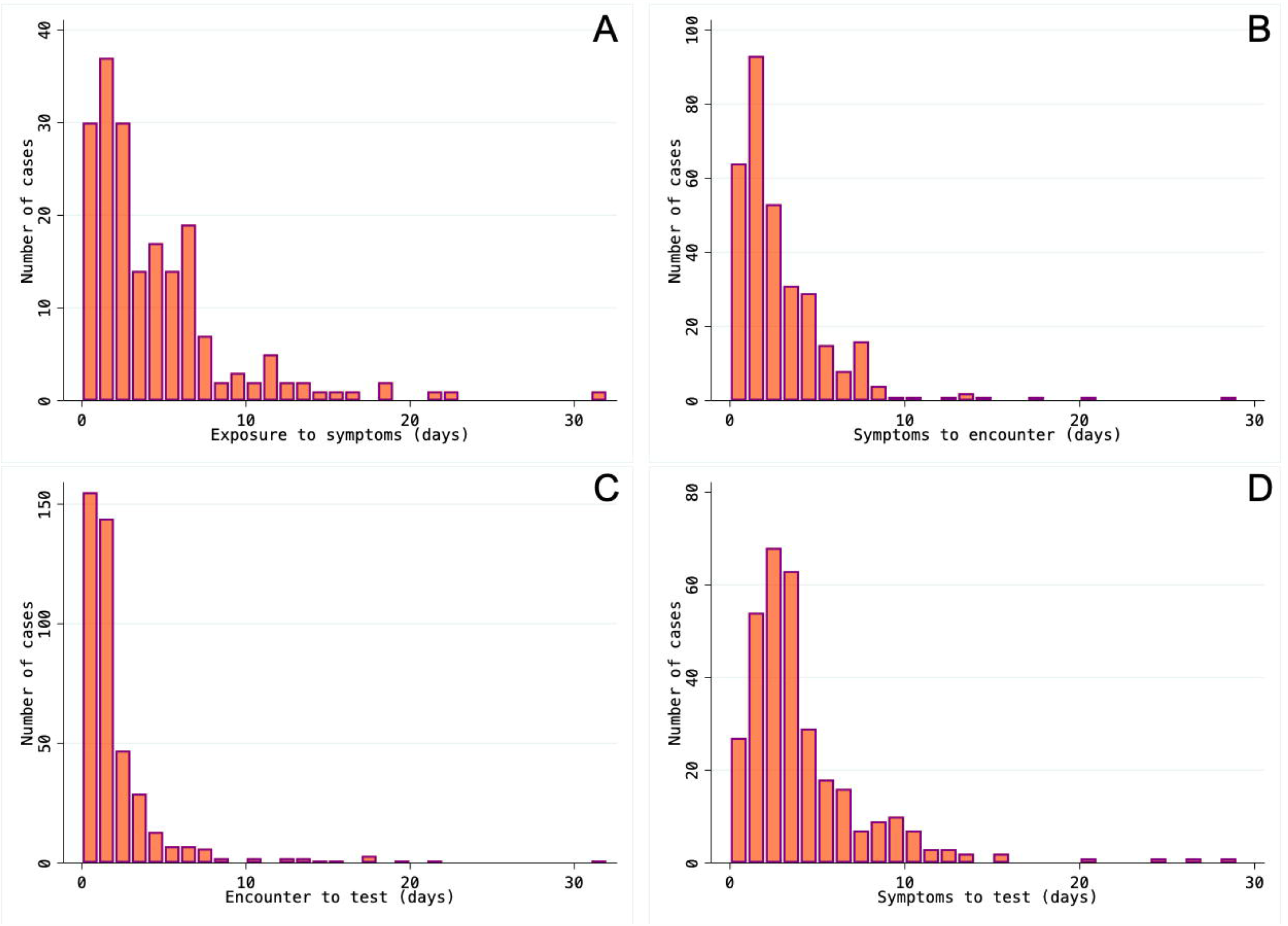
Histogram of timeline distribution. Histogram of days from exposure to symptoms (A), from symptom onset to initial presentation to care (B), from presentation to care to test (C) and from symptom onset to test (D). In (A), the patients with greater than 30 days since exposure presented with MIS-C.

### Symptoms

Approximately 79% of patients presented with symptoms, the most common being fever (42.5%), cough (37.0%), rhinorrhea (30.7%), pharyngitis (20.3%), headache (18.6%), fatigue (13.4%), myalgias (13.2%), and ageusia (11.6%) (**Table 3**). No patients reported chilblains or “COVID-toes.” When evaluated by age, those in the youngest group (0-2 years) were more likely to present with fever, cough, and rhinorrhea than older children (p<0.005) (**Figure 4**). Older children were more likely to have more numerous symptoms (p=0.001). Twenty-one percent of cases in the cohort were asymptomatic and were identified through testing after a known exposure (70.5%), pre-procedural screening at CHP (15.9%), or via testing required to return to school or sports (10.3%) (**Supplementary Table 2**).

**Table 3:**
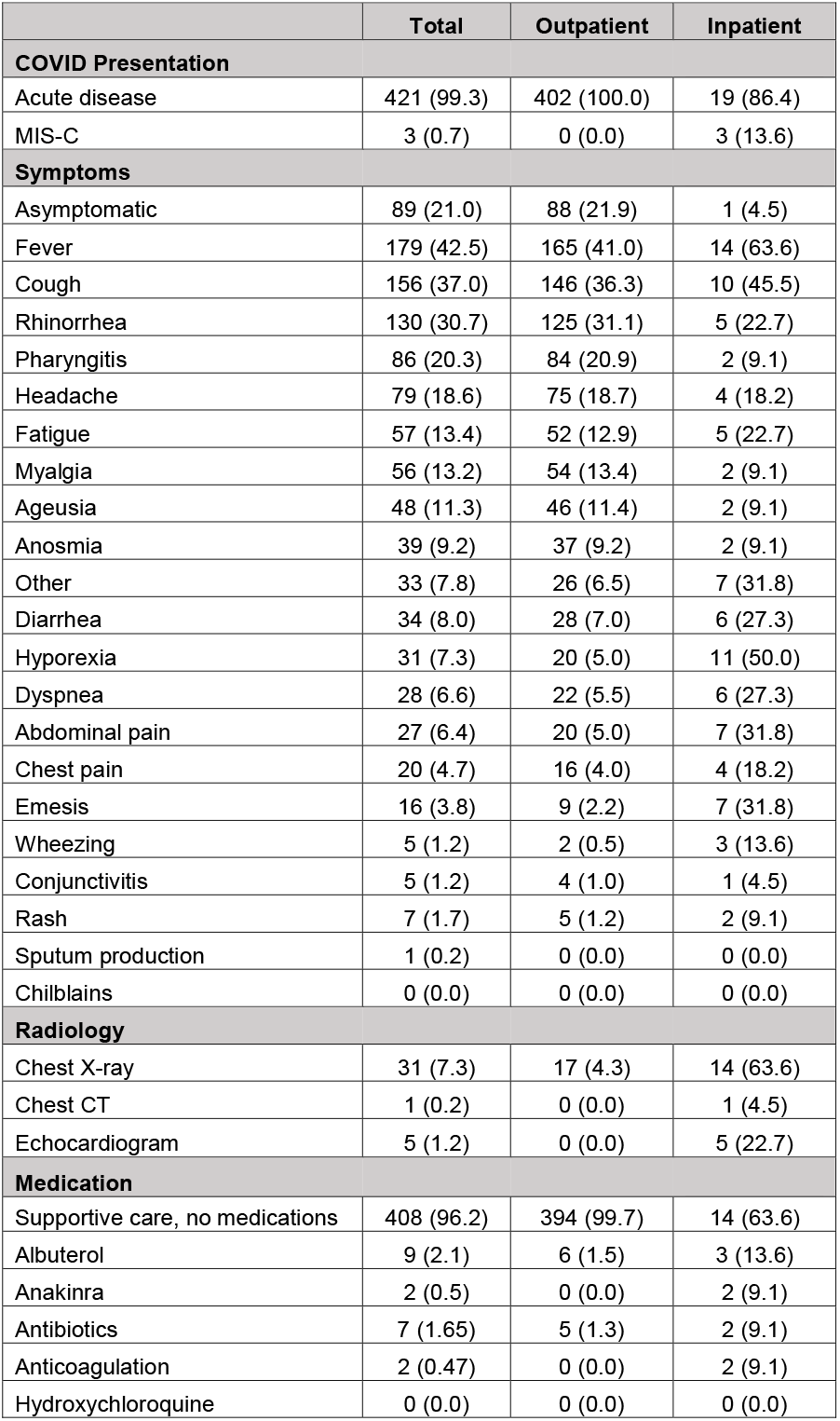
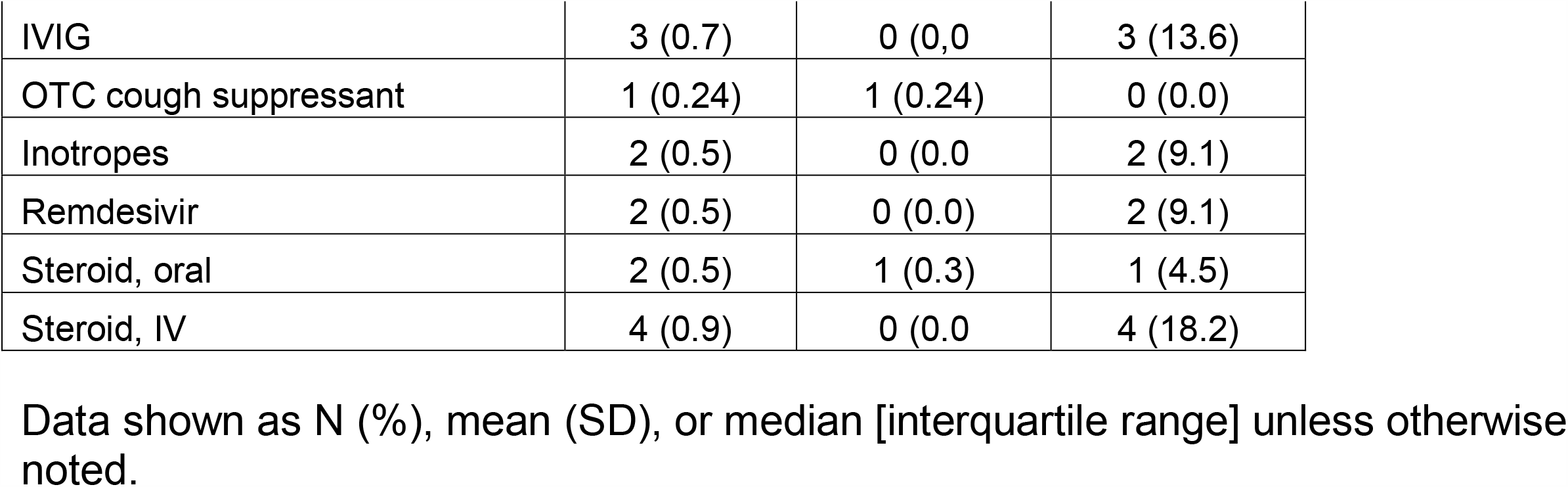
Disease Presentation.

### Severe disease

Most cases with acute COVID-19 were treated as outpatients and recovered at home (95.4%), but 22 required hospital admission: nineteen with acute disease and three with MIS-C. Fifteen patients were hospitalized for management of acute COVID-19 symptoms, while the others were admitted for other medical problems and were found to be positive during evaluation. Patients with chronic medical conditions were more likely to be hospitalized than those without significant past medical history (10.8% vs 1.9%, p<0.001). Hospitalized patients were significantly younger than those followed as outpatients (7.2 [7.4] vs 12.8 [7.0] years, p<0.001), with the youngest age group representing 45.5% of hospitalizations (**Table 4**). Black/Hispanic patients were more 5.8 times more likely to be admitted than non-Hispanic white patients (12.0% vs 2.9%, respectively; OR=5.8 [95%CI=1.5-22.8] after adjusting for age, chronic conditions, and the number of days between symptom onset and testing; p=0.012). Hospitalized patients were more likely to present with fever, dyspnea, wheezing, chest pain, hyporexia, vomiting, abdominal pain, or diarrhea (p<0.05).

**Table 4:** Hospitalized patients.

|  | Acute COVID-19 | MIS-C |
| --- | --- | --- |
| **Location** |  |  |
| Admitted Patients | 19 | 3 |
| Admitted for COVID | 15 (78.9) |  |
| Admitted for other reasons | 4 (21.1) |  |
| Floor | 17 (89.5) | 0 |
| LOS, days | 2.2 (2.9) | n/a |
| PICU | 2 (10.5) | 3 (100.0) |
| LOS, days | 6.5 (0.7) | 11.3 (4.0) |
| **Demographics** |  |  |
| **Sex** |  |  |
| Female | 8 (42.1) | 1 (33.3) |
| Male | 11 (57.9) | 2 (66.7) |
| **Age, years** |  |  |
| Mean (SD) | 6.7 (8.1) | 8.8 (1.4) |
| Median [IQR] | 1.3 [0.4-15.3] | 8.7 [7.5-10.3] |
| **Race/ ethnicity<sup>1</sup>** |  |  |
| White | 8 (42.1) | 1 (33.3) |
| Black | 9 (47.4) | 2 (66.7) |
| Asian | 0 (0.0) | 0 (0.0) |
| AIAN | 0 (0.0) | 0 (0.0) |
| Other | 1 (5.3) | 0 (0.0) |
| Hispanic/Latino | 1 (5.3) | 1 (33.3) |
| **Underlying condition** |  |  |
| No | 5 (26.3) | 3 (100.0) |
| Yes | 14 (73.7) | 0 (0.0) |
| Reported home medications | 9 (47.4) | 0 (0.0) |
| **Radiology** |  |  |
| Chest X-ray | 11 (57.9) | 3 (100.0) |
| - Abnormal | 5 (45.5) | 0 (0.0) |
| Chest CT | 1 (5.3) | 0 (0.0) |
| - Abnormal | 1 (100.0) | 0 (0.0) |
| Echocardiogram | 2 (10.5) | 3 (100.0) |
| - Abnormal | 0 (0.0) | 1 (33.3) |
| **Management** |  |  |
| Supportive care, no medications | 14 (73.7) | 0 (0.0) |
| Albuterol | 3 (15.8) | 0 (0.0) |
| Anakinra | 0 (0.0) | 0 (0.0) |
| Antibiotics | 1 (5.3) | 2 (66.7) |
| Anticoagulation | 0 (0.0) | 2 (66.7) |
| Hydroxychloroquine | 0 (0.0) | 3 (100.0) |
| IVIG | 0 (0.0) | 1 (33.3) |
| Inotropes | 0 (0.0) | 2 (66.7) |
| Remdesivir | 1 (5.3) | 0 (0.0) |
| Steroid, Oral | 1 (5.3) | 1 (33.3) |
| Steroid, IV | 1 (5.3) | 2 (66.7) |
| **Maximum Respiratory Support** |  |  |
| Room Air | 16 | 0 (0.0) |
| Nasal Cannula | 1 (5.3) | 1 (33.3) |
| BiPAP | 2 (10.6) | 1 (33.3) |
| Mechanical Ventilation | 0 (0.0) | 1 (33.3) |
| **Inflammatory Markers** |  |  |
| Patients with any lab | 6 (31.6) | 3 (100.0) |
| CRP, mg/dL | 6.7 (5.4) | 26.1 (11.2) |
| Procalcitonin, ng/mL | n/a | 49.2 (2.9) |
| Ferritin, ng/mL | 261.0 (84.4) | 936.9 (155.9) |
| BNP, pg/mL | 801 (n=1) | 1403 (1228) |
| Troponin, ng/mL | 0.252 (n=1) | 0.7 (1.0) |
| **Outcome** |  |  |
| Discharged | 19 (100.0) | 3 (100.0) |
| Died | 0 (0.0) | 0 (0.0) |

Data shown as N (%), mean (SD), or median [interquartile range] unless otherwise noted. <sup>1</sup>Allowed to choose more than one response.

Among hospitalized patients, five were admitted to the PICU; two for acute disease requiring BiPAP and three for MIS-C. There were no differences in age between PICU and non-PICU patients (p=0.11), but all five PICU cases were Black or Hispanic. Average length of stay (LOS) was 2.2 (2.9) days for patients admitted to the floor vs. 9.4 (3.9) days for PICU patients (p<0.001).

Most patients admitted received radiographic studies, including chest X-ray (11), chest CT (1), or echocardiograms (2). Five chest X-rays were reported as abnormal and showed increased interstitial markings (4), lobar consolidation (1), and/or multifocal opacities (1). Treatment included supportive care (14), albuterol (3), and one patient each received remdesivir, IV steroids, oral steroids, and empiric antibiotics (**Table 4**). All patients were discharged home and there were zero deaths.

### MIS-C

Three patients during the study period were admitted to the PICU for MIS-C, with an average length of stay of 11.3 (4.0) days (range 9-16 days) (**Table 4**). Two patients were male; two identified as Black and one as Hispanic white. Mean age was 8.8 (1.4) years (range 7.5-10.3 years). All were otherwise previously healthy. All had normal chest X-rays, and all received echocardiograms, of which one demonstrated ventricular dysfunction with coronary dilatation. All MIS-C patients were treated with IV steroids and immunoglobulin; one received remdesivir and two received anakinra. Two required inotropic support. All patients were ultimately discharged home and there were no deaths.

## DISCUSSION

In this report, we provide a comprehensive clinical description of pediatric COVID-19 in Western PA, a region with a low to moderate case burden. We describe clinical presentation and healthcare utilization for the breadth of pediatric disease, ranging from asymptomatic patients identified on preprocedural screening to critically ill patients with the presentation of either acute COVID-19 or MIS-C. Due to the market capture of the participating institutions, we are able to provide a sampling of cases that likely mirrors all pediatric disease in the area, with a bias toward inclusion of children who require hospitalization or intensive care. These cases span the date range of March to August 2020, a period in which all schools were closed to in-person learning in our area.

The demographics of our cohort mirror some national trends, demonstrating that Black and Hispanic adults ^17,18,19^ and children ^20,21^ are more likely to be infected with or have worse outcomes from COVID-19. According to the most recent available census data^22^ the largest ethnic groups in Allegheny County, PA, are non-Hispanic whites (79.9%) followed by non-Hispanic Blacks (13.4%). While our registry encompasses an entire region of the state and not solely this county, Black patients are overrepresented among infected children (24.5% of patients in the registry). While the total number is small, all individuals who presented with MIS-C were from minority groups, which also aligns with prior reports^23,24^. Racial/ethnic differences in hospitalization risk remained significant after accounting for age, sex, the presence of chronic conditions, and the time lapse between symptom onset and COVID-19 testing.

This study also highlights, as is true for many pediatric conditions, that symptoms of SARS-CoV-2 infection vary with the age of the child. The youngest children were more likely to present with fever, cough, and rhinorrhea than older children, who more frequently presented with a syndrome of more numerous symptoms that may not include fever. Evaluating children, particularly in the outpatient setting, through this lens can be beneficial in recognizing potential SARS-CoV-2 infection, particularly as it presents with symptoms similar to other respiratory viral pathogens^25^.

Based on the data in our area, we estimate a prevalence of MIS-C of approximately 2.0 per 1,000 SARS-CoV-2 pediatric infections. However, this is likely an overestimate, since children can have asymptomatic disease (up to 2.2% of well-children based on all comers for pre-procedure screening^26^), and thus symptom-based screening likely excludes some infected patients. In addition, our center receives transfers for more severe disease in the region, including MIS-C. Other estimates of MIS-C prevalence have ranged from a nationwide rate of 2.0 per 1,000 (694/355,123)^27^ to the PA statewide rate of 4.3 per 1,000 (43/10,011)^16^.

Another important observation during this study is the rapid rise in the rate of telehealth appointments. While CCP did not routinely use telemedicine prior to this pandemic, they rapidly adapted to being able to provide care and counseling (especially for patients with respiratory symptoms), monitor patients, and arrange for SARS-CoV-2 testing at an off-site drive-through testing facility to decrease exposure risk in their offices. Several subspecialty disciplines also moved the majority of routine outpatient encounters to virtual visits within days of the first case detected in the region. Telehealth will continue to play an important role in pediatric healthcare delivery as we move into the winter respiratory illness season, when SARS-CoV-2 will be circulating in addition to other typical winter viruses such as influenza and RSV. This will also be crucial given the additional burden placed on outpatient healthcare providers who are being asked to provide clearance for ill children to return to school in the era of COVID-19.

This study has several strengths. Our registry is based on information abstracted directly from the EHR by providers involved in the care of these children, which provides more detailed and accurate information than reports based on ICD-10 codes or claims data. We were therefore able to break down patterns by age group. The registry cohort represents a wide range of SARS-CoV-2 pediatric clinical presentations outside of a pandemic “hotspot”; disease severity is thus less likely affected by strain on the system or difficulties accessing care. Our results advance prior pediatric literature showing higher risk of SARS-CoV-2 infection in minority children to show that these children are also at higher risk of severe disease, as has been reported in adults. At the same time, we must acknowledge study limitations. First, given the scope of practice at our referral center, we cannot rule out bias towards more severe disease; however, most cases were detected and followed as outpatients at CCP. Second, the current report does not yet include long-term patient outcomes and, thus, these data provide a limited understanding of patient recovery or long-term sequelae. However, this real-time registry will allow for additional follow-up on these metrics. We can use this tool to measure how dynamics of the outbreak in the pediatric population change with added factors in the coming months, including changes in child behavior with the school year, the respiratory virus season, and hopefully the eventual introduction of vaccines.

In summary, this report provides a comprehensive overview of the breadth of presentation of pediatric infection with SARS-CoV-2 in a single geographic area. In general, pediatric patients tolerate this infection well, with only a small proportion (∼5%) requiring hospital admission, and even fewer (∼1.2%) presenting with critical illness. Large, multicenter collaborative efforts will be crucial to better understand factors associated with severe pediatric SARS-CoV-2 disease, particularly in racial and ethnic minorities at higher risk.

## Data Availability

Additional data available upon request.

## Acknowledgements

The authors would like to acknowledge the physicians, advanced practice providers, nurses, and other healthcare professionals on the front line of the COVID-19 pandemic at UPMC CHP and CCP; the Allegheny County Health Department and the PA Department of Health; and the leadership at UPMC CHP and the University of Pittsburgh Schools of Medicine and Public Health.

## Funding

This work was supported in part by the Clinical and Translational Science Institute, University of Pittsburgh supported by the National Institutes of Health/National Center for Advancing Translational Sciences [grant number UL1-TR-001857], the Pediatric Infectious Disease Society and St. Jude Children’s Research Hospital Fellowship Program in Basic and Translational Research to MCF, the National Institutes of Health/ National Heart, Lung, and Blood Institute [grant number T32-HL129949] to KMG, the Pediatric Scientist Development Program (PSDP) sponsored by the Association of Medical School Pediatric Department Chairs (AMSPDC) funded by the National Institutes of Health/ National Institute of Child Health and Human Development [grant K12HD000850-34] to GJR, and the National Institute of Health/National Heart, Lung, and Blood Institute [grant number R01-HL149693] to EF. The authors do not have any conflicts of interest to declare related to this work.

## Notes

### Competing Interest Statement

The authors have declared no competing interest.

### Author Declarations

Patient information was securely handled as approved by the institutional review board at the University of Pittsburgh (Protocol # STUDY20030250). All data were deidentified prior to analysis.

## References

1. Zhu N, Zhang D, Wang W, et al. A Novel Coronavirus from Patients with Pneumonia in China, 2019. New Engl J Medicine. Published online 2020. doi:10.1056/nejmoa2001017

2. Department ACH. Allegheny County Health Department COVID-19 Dashboard. https://www.alleghenycounty.us/Health-Department/Resources/COVID-19/COVID-19.aspx.

3. Parri N, Lenge M, Buonsenso D, Group CI in PED (CONFIDENCE) R. Children with Covid-19 in Pediatric Emergency Departments in Italy. New Engl J Med. 2020;383(2):187–190. doi:10.1056/nejmc2007617

4. Lu X, Zhang L, Du H, et al. SARS-CoV-2 Infection in Children. New Engl J Med. 2020;382(17):1663–1665. doi:10.1056/nejmc2005073

5. Dong Y, Mo X, Hu Y, et al. Epidemiology of COVID-19 Among Children in China. Pediatrics. 2020;145(6):e20200702. doi:10.1542/peds.2020-0702

6. Team CC-19 R, Bialek S, Gierke R, et al. Coronavirus Disease 2019 in Children - United States, February 12-April 2, 2020. Mmwr Morbidity Mortal Wkly Rep. 2020;69(14):422–426. doi:10.15585/mmwr.mm6914e4

7. Feldstein LR, Rose EB, Horwitz SM, et al. Multisystem Inflammatory Syndrome in U.S. Children and Adolescents. New Engl J Med. 2020;383(4):334–346. doi:10.1056/nejmoa2021680

8. Toubiana J, Poirault C, Corsia A, et al. Kawasaki-like multisystem inflammatory syndrome in children during the covid-19 pandemic in Paris, France: prospective observational study. Bmj. 2020;369:m2094. doi:10.1136/bmj.m2094

9. Cheung EW, Zachariah P, Gorelik M, et al. Multisystem Inflammatory Syndrome Related to COVID-19 in Previously Healthy Children and Adolescents in New York City. Jama. 2020;324(3):294. doi:10.1001/jama.2020.10374

10. Ramcharan T, Nolan O, Lai CY, et al. Paediatric Inflammatory Multisystem Syndrome: Temporally Associated with SARS-CoV-2 (PIMS-TS): Cardiac Features, Management and Short-Term Outcomes at a UK Tertiary Paediatric Hospital. Pediatr Cardiol. Published online 2020:1–11. doi:10.1007/s00246-020-02391-2

11. Hoang A, Chorath K, Moreira A, et al. COVID-19 in 7780 pediatric patients: A systematic review. Eclinicalmedicine. 2020;24:100433. doi:10.1016/j.eclinm.2020.100433

12. Guo C-X, He L, Yin J-Y, et al. Epidemiological and clinical features of pediatric COVID-19. Bmc Med. 2020;18(1):250. doi:10.1186/s12916-020-01719-2

13. Zhen-Dong Y, Gao-Jun Z, Run-Ming J, et al. Clinical and Transmission Dynamics Characteristics of 406 Children with Coronavirus Disease 2019 in China: A Review. J Infection. 2020;81(2):e11–e15. doi:10.1016/j.jinf.2020.04.030

14. Barney A, Buckelew S, Mesheriakova V, Raymond-Flesch M. The COVID-19 Pandemic and Rapid Implementation of Adolescent and Young Adult Telemedicine: Challenges and Opportunities for Innovation. J Adolescent Health. 2020;67(2):164–171. doi:10.1016/j.jadohealth.2020.05.006

15. Harris PA, Taylor R, Thielke R, Payne J, Gonzalez N, Conde JG. Research electronic data capture (REDCap)--a metadata-driven methodology and workflow process for providing translational research informatics support. Journal of biomedical informatics. 2009;42(2):377–381. doi:10.1016/j.jbi.2008.08.010

16. Health PD of. Pennsylvania Department of Health Coronavirus (COVID-19). Published 2020. accessed September 15, 2020. https://www.health.pa.gov/topics/disease/coronavirus/Pages/Coronavirus.aspx

17. Golestaneh L, Neugarten J, Fisher M, et al. The association of race and COVID-19 mortality. Eclinicalmedicine. Published online 2020:100455. doi:10.1016/j.eclinm.2020.100455

18. Price-Haywood EG, Burton J, Fort D, Seoane L. Hospitalization and Mortality among Black Patients and White Patients with Covid-19. New Engl J Med. 2020;382(26):2534–2543. doi:10.1056/nejmsa2011686

19. Bandi S, Nevid MZ, Mahdavinia M. African American children are at higher risk of COVIDLJ19 infection. Pediatr Allergy Immu. Published online 2020. doi:10.1111/pai.13298

20. Kim L, Whitaker M, O’Halloran A, et al. Hospitalization Rates and Characteristics of Children Aged <18 Years Hospitalized with Laboratory-Confirmed COVID-19 — COVID-NET, 14 States, March 1–July 25, 2020. Morbidity Mortal Wkly Rep. 2020;69(32):1081–1088. doi:10.15585/mmwr.mm6932e3

21. Goyal MK, Simpson JN, Boyle MD, et al. Racial/Ethnic and Socioeconomic Disparities of SARS-CoV-2 Infection Among Children. Pediatrics. 2020;146(4):e2020009951. doi:10.1542/peds.2020-009951

22. Bureau USC. QuickFacts County Search. Published 2019. accessed September 3, 2020. https://www.census.gov/quickfacts/alleghenycountypennsylvania

23. Davies P, Evans C, Kanthimathinathan HK, et al. Intensive care admissions of children with paediatric inflammatory multisystem syndrome temporally associated with SARS-CoV-2 (PIMS-TS) in the UK: a multicentre observational study. Lancet Child Adolesc Heal. 2020;4(9):669–677. doi:10.1016/s2352-4642(20)30215-7

24. Riphagen S, Gomez X, Gonzalez-Martinez C, Wilkinson N, Theocharis P. Hyperinflammatory shock in children during COVID-19 pandemic. Lancet. 2020;395(10237):1607–1608. doi:10.1016/s0140-6736(20)31094-1

25. Yonker LM, Neilan AM, Bartsch Y, et al. Pediatric SARS-CoV-2: Clinical Presentation, Infectivity, and Immune Responses. J Pediatrics. Published online 2020. doi:10.1016/j.jpeds.2020.08.037

26. Sola AM, David AP, Rosbe KW, Baba A, Ramirez-Avila L, Chan DK. Prevalence of SARS-CoV-2 Infection in Children Without Symptoms of Coronavirus Disease 2019. Jama Pediatr. 2021;175(2). doi:10.1001/jamapediatrics.2020.4095

27. Prevention C for DC and. CDC COVID Data Tracker. Published 3AD. accessed September 3, 2020. https://covid.cdc.gov/covid-data-tracker/?CDC_AA_refVal=https%3A%2F%2Fwww.cdc.gov%2Fcoronavirus%2F2019-ncov%2Fcases-updates%2Fcases-in-us.html#demographics

