## Supplemental Table 1 for "A Comprehensive Clinical Description of Pediatric SARS-CoV-2 Infection in Western Pennsylvania"

**Supplementary Table 1: Chronic conditions in pediatric COVID-19 cases**

|  | **Outpatient** | **Inpatient** |
| --- | --- | --- |
| **EHR ICD-10 codes of positive patients** | **Frequency (%)** | **Frequency (%)** |
| Asthma | 45 (10.6) | 2 (9.1) |
| Hyperkinetic disorders (e.g. ADHD) | 34 (8.0) | 1 (4.5) |
| Other anxiety disorders | 17 (4.0) | 1 (4.5) |
| Vasomotor and allergic rhinitis | 15 (3.5) | --- |
| Congenital malformations of cardiac septa | 8 (1.9) | 1 (4.5) |
| Pervasive developmental disorders | 8 (1.9) | 1 (4.5) |
| Migraine | 7 (1.7) | --- |
| Obesity | 7 (1.7) | 4 (18.2) |
| Atopic dermatitis | 6 (1.4) | --- |
| Depressive episode | 5 (1.2) | --- |
| Type 1 diabetes mellitus | 4 (0.9) | 2 (9.0) |
| Acne | 3 (0.7) | --- |
| Convulsions, not elsewhere classified | 3 (0.7) | --- |
| Epilepsy | 3 (0.7) | 1 (4.5) |
| Excessive, frequent and irregular menstruation | 3 (0.7) | --- |
| Gastroesophageal reflux disease | 3 (0.7) | --- |
| Irritable bowel syndrome | 3 (0.7) | 1 (4.5) |
| Other coagulation defects | 3 (0.7) | --- |
| Other functional intestinal disorders | 3 (0.7) | --- |
| Pain and other conditions associated with female genital organs and menstrual cycle | 3 (0.7) | --- |
| Atresia of bile ducts | 1 (0.2) | 1 (4.5) |
| Craniosynostosis | 1 (0.2) | 1 (4.5) |
| Cystic fibrosis | 1 (0.2) | 1 (4.5) |
| Diffuse brain injury | 1 (0.2) | 1 (4.5) |
| Double outlet right ventricle | 1 (0.2) | 2 (9.0) |
| Hypospadias | 1 (0.2) | 1 (4.5) |
| Malignant neoplasm of adrenal gland | 1 (0.2) | 1 (4.5) |
| Other congenital malformations of nervous system | 1 (0.2) | --- |
| Other diseases of gallbladder | 1 (0.2) | --- |
| Other diseases of liver | 1 (0.2) | --- |
| Other disorders of Eustachian tube | 1 (0.2) | --- |
| Other perinatal digestive system disorders | 1 (0.2) | 1 (4.5) |
| Other sex chromosome abnormalities, male phenotype, not elsewhere classified | 1 (0.2) | --- |
| Paroxysmal tachycardia | 1 (0.2) | --- |
| Personal history of other diseases and conditions | 1 (0.2) | --- |
| Phacomatosis, not elsewhere classified | 1 (0.2) | --- |
| Reaction to severe stress, and adjustment disorders | 1 (0.2) | --- |
| Relapsing fevers | 1 (0.2) | 1 (4.5) |
| Renal agenesis and other reduction defects of kidney | 1 (0.2) | --- |
| Scoliosis | 1 (0.2) | --- |
| Seborrheic dermatitis | 1 (0.2) | --- |
| Specific developmental disorders of scholastic skills | 1 (0.2) | --- |
| Specific developmental disorders of speech and language | 1 (0.2) | --- |
| Spina bifida | 1 (0.2) | --- |
| Symptoms and signs concerning food and fluid intake | 1 (0.2) | --- |
| Transplanted organ and tissue status | 1 (0.2) | 1 (4.5) |
| Undescended testicle | 1 (0.2) | --- |

ICD-10 codes abstracted from problem list in the EHR.
