## Supplemental Table 2 for "A Comprehensive Clinical Description of Pediatric SARS-CoV-2 Infection in Western Pennsylvania"

**Supplementary Table 2: Reported exposure in asymptomatic patients**

| **Why tested?** | **Cases (%)*** |
| --- | --- |
| Known exposure | 62 (70.5) |
| Pre-procedure screening | 14 (15.9) |
| Required for school | 7 (8.0) |
| Required for sports | 2 (2.3) |
| Unclear / not recorded | 3 (3.4) |
| TOTAL | 88 |
